# Altered neutrophil G-protein receptor signalling linked to impaired chemotaxis and increased ROS and NET production in older people with frailty

**DOI:** 10.64898/2025.12.16.25342352

**Authors:** Genna Ali Abdullah, Jack Walsh, Andrew Sellin, Cody McLean, Asangaedem Akpan, Marie M Phelan, Helen L Wright

**Author notes:** Corresponding author details: Dr Helen L Wright, University of Liverpool, Institute of Life Course and Medical Sciences, William Henry Duncan Building, 6 West Derby Street, Liverpool, L7 8TX.

## Abstract

**Background:** Immune function alters with age, and is often accompanied by chronic, low-grade inflammation (inflammageing). In individuals with frailty, inflammageing is increased, reducing immune function and increasing susceptibility to serious outcomes from infection. In this study we investigated the changes that take place in human neutrophils during healthy ageing and ageing with frailty (FR) using RNAseq and functional assays. We also compared neutrophil phenotype in frailty with rheumatoid arthritis (RA) neutrophils.

**Methods:** Blood neutrophil RNAseq data were analysed using IDEP2 and Ingenuity Pathway Analysis (IPA). Neutrophil phenotype was assessed for reactive oxygen species (ROS) production, neutrophil extracellular trap (NET) release, chemotaxis, and bacterial killing capacity. Experimental data were analysed by ANOVA in R (v4.5.1).

**Results:** RNAseq identified activation of G-protein coupled receptors, interferon and cytokine receptor signalling, and chemotaxis pathways in frail individuals (n=10) compared to healthy older (n=9) and healthy younger people (n=8, adj. *p*<0.05). FR neutrophils expressed more IL-8 receptors (CXCL1 and CXCL2) and CD177 on their surface (*p*<0.05). FR neutrophil phenotype was similar to RA, with significantly higher ROS production and impaired chemotaxis and bacterial killing capacity (*p*<0.05). FR neutrophils also released significantly more NETs in response to LPS (10ng/mL, *p*<0.05).

**Conclusions:** This work provides novel insight into the changes to neutrophil phenotype associated with ageing in good health and ageing with frailty, and highlights similarities between inflammageing in frailty and chronic inflammation in RA. This may be important in the development of future therapeutics and/or health management strategies to support healthy living as we age.

**Graphical Abstract:** In this study we have combined RNAseq with functional experiments to describe the phenotype of neutrophils from people with frailty. We found that frailty neutrophils had altered gene expression with activation of G-protein coupled receptors, interferon and cytokine receptor signalling, and chemotaxis pathways. This was associated with altered ROS and NET production, impaired chemotaxis and impaired bacterial killing capacity compared to healthy older neutrophils. Created in BioRender. Abdullah, G. (2026) https://BioRender.com/ze634lt and https://BioRender.com/7khpj2z.

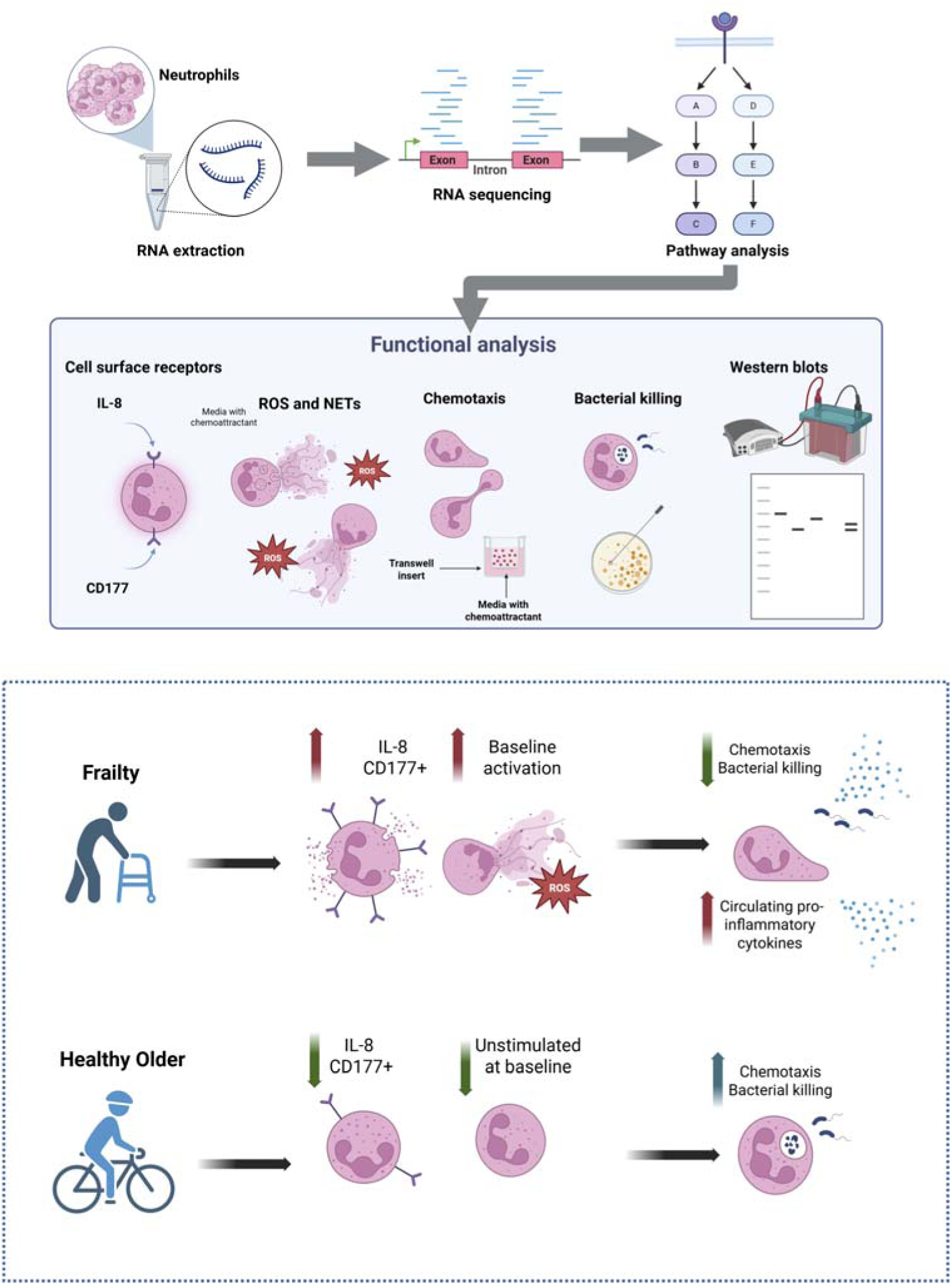

## Introduction

Neutrophils are effector innate immune cells that play a key role in immune responses to microorganisms. When activated, neutrophils have the capacity to phagocytose and kill particulate pathogens such as bacteria and fungi, release a range of cytotoxic molecules including reactive oxygen species (ROS), proteases, and neutrophil extracellular traps (NETs), as well as produce inflammatory cytokines and chemokines to orchestrate the inflammatory response [1]. There are limited primary studies on how neutrophil function changes during healthy human ageing [2]. It is established that ageing is heterogeneous, and there is currently a huge gap in the literature regarding neutrophil function during healthy ageing and within people with frailty in the absence of acute infections. There are mixed reports from mouse ageing models, with reports of chemotaxis being reduced in older age and a decrease in NET production in response to LPS from *Staphylococcus aureus* compared to young mice [3]. A hyperreactive systemic inflammatory response in aged mice with increased levels of MPO in response to LPS from *Pseudomonas aeruginosa* has also been described [4]. These differences probably reflect various factors, including the characteristics of genetically homogeneous inbred mouse strains [5].

In humans, neutrophils are activated by many self and non-self-compounds which elicits their classic functions. There are some reports that neutrophils display an age-related impairment in their effector functions. However, the experimental evidence of neutrophil function in frailty without an infection is virtually non-existent, with only one previous publication which measured chemotaxis [6]. Previously, several immune functions such as neutrophil and macrophage chemotaxis, phagocytosis, natural killer (NK) cell activity, and lymphoproliferation were reported as predictive markers of biological age, and although showing an age-related decline, they are preserved in long-lived individuals [7].

Transcriptomic activity occurs in neutrophils during development and maturity in the bone marrow [8] and expands rapidly during activation in the blood and tissues to regulate neutrophil functions [9]. It is now over 20 years since initial studies demonstrated that neutrophils are capable of initiating gene expression in response to infectious stimuli [10]. The unfolding of metabolic plasticity in neutrophils has also supported these reports of transcriptional adaptability. These reports may be unsurprising since it is now recognised that neutrophils generate and release cytokines, increase their transcriptional activity during NET production, as well as the ability of ROS to activate transcription factors involved in DNA repair [11].

To date, there have been differences reported in the neutrophil genes upregulated during autoimmune/chronic inflammatory conditions [12–15] compared to pathogenic infection with bacteria [16, 17], fungi [18, 19] and viruses [20–23]. Within the last decade, the latest single cell analysis techniques have revealed distinct populations, even amongst mature neutrophils [24]. As expected, there are crossovers with regard to the genes that are upregulated in neutrophils in response to pathogens in general, but it is now evident that there is not a “one size fits all” approach from neutrophils in response to stimuli from different inflammatory agents [25]. Instead, neutrophils display exceptional adaptability from their transcriptome through to their functional responses to microenvironmental cues. This newfound appreciation of neutrophil heterogeneity and plasticity has profound implications for understanding their roles in health and disease.

There are currently no studies reporting the differences in gene expression between people with frailty compared to healthy older individuals in the absence of infection, or those genes altered between healthy older and healthy younger people. This is the first study to explore the differences between unstimulated circulating blood neutrophils from people of different ages, to investigate the effects of healthy ageing and ageing with frailty on neutrophil transcriptomics and to validate the changes in gene expression with a range of functional experiments to explore pathogenic neutrophil functions in healthy ageing and in frailty. An inflammatory control group of people with rheumatoid arthritis (RA) has also been included in the study design to test our hypothesis that frailty has an underlying inflammatory phenotype. We show that neutrophils from people with frailty have an activated transcriptome, predicting altered cell signalling and dysregulated host defence functions. We then demonstrate experimentally that neutrophils have altered phenotype in people with frailty, and show similarities in dysregulated ROS and NET production, migration capacity and bacterial killing ability with inflammatory neutrophils from people with RA.

## Methods

### Study Population

All participants with frailty fulfilled the Rockwood Clinical Frailty Scale criteria [26] for the diagnosis of frailty (score ≥5). All participants with rheumatoid arthritis fulfilled the American College of Rheumatology 2010 criteria [27]. Participant demographics are shown in Supplementary Table 1. Healthy older people were aged 65 years or over. Healthy young participants were aged 18-25 years. All participants were free of infection for at least 4 weeks prior to participating in the study.

### Neutrophil Isolation

Neutrophils were isolated from peripheral blood using Ficoll Paque or the Stem Cell human neutrophil isolation kit within 1h of venipuncture as described previously [28]. Erythrocytes were removed by hypotonic lysis in ammonium chloride buffer for 3min. Neutrophils were resuspended in RMPI 1640 media (+L-glutamine) and concentration adjusted to 5×10^6^cells/ml. Purity was assessed by cytospin and found to be routinely >95-99%.

### RNA Extraction

RNA was extracted from 5×10^6^ neutrophil using Trizol and chloroform as previously described [29]. Total RNA was cleaned using the Qiagen RNeasy kit, including the on-column DNase digestion. RNA was quantified using nanodrop2000, snap frozen in liquid nitrogen and then stored at -80°C.

### RNA Sequencing

RNA integrity was measured using the Agilent 2100 fragment analyser. Samples with RNA integrity (RIN)>7.0 passed QC and were processed for RNA sequencing. Total RNA was enriched with poly-A selection and paired-end cDNA libraries was generated for each sample using standard protocols. RNA sequencing was performed using the DNBseq platform by BGI, Hong Kong (https://www.bgi.com). Sequencing generated 100 base pair (bp) paired end reads for each sample. Raw reads were mapped to the human genome (hg38) using HISAT2 [30]. Raw gene counts were generated by featureCounts [31]. Differentially expressed genes (DEGs) were identified using DESeq2 using a negative binomial generalized linear model (GLM) within the IDEP2 web-based software (https://bioinformatics.sdstate.edu/idep/) [32]. DEGs were considered with a false discovery rate (FDR)-adjusted *p*-value threshold of <0.05 and a 1.5-fold change in increased or decreased expression. Multivariate principal component analysis (PCA) and heatmaps were produced from variable stabilising transformation (VST) count data to explore data dimensionality and structure using R (v4.5.1).

### Bioinformatics Analyses

DEGs between sample groups were selected for further analysis. Pathway enrichment analysis of DEGs was performed via the web-based tool Integrated Differential Expression & Pathway analysis (IDEP) software [32] using gene set enrichment analysis (GSEA) from the Gene Ontology Biological Pathways (GOBP). Kyoto Encyclopedia of Genes and Genomes (KEGG), and REACTOME pathway databases. Ingenuity Pathway Analysis software (IPA; QIAGEN, Valencia, CA, https://www.qiagenbioinformatics.com/products/ingenuity-pathway-analysis) was also used to perform canonical pathway enrichment analysis, upstream regulator prediction and to perform reconstruction of gene networks. Statistical significance was adjusted using the Benjamini-Hochberg method, with an adjusted p-value threshold of <0.05.

### Western Blotting

Neutrophils (5×10^6^/mL) were centrifuged at 500g for 3 min and lysed in 1mL boiling Laemmli buffer containing 100mM dithiothreitol (DTT). Protein lysates were boiled at 100°C for 3min before loading 10µL of each sample into 10% 1mm bis-tris gels. Gels were run using MOPs SDS running buffer (Merck, MPM0PS) at 180V for 31min before transferring onto PVDF membranes in sandwich cassettes using bis-tris running buffer (Merck, MPTRB) at 100V for 60min on ice. Membranes were blocked with 5% non-fat dry milk powder in Tris buffer for 1hr before incubating with 1:1000 primary antibodies overnight at 4°C. Membranes were then washed 3x for 5min each time with wash buffer (1x TBS, 0.075% Tween20) and incubated with secondary antibodies (1:5000) for 1h at room temperature on a platform shaker before washing for 5min 3x with wash buffer. Membranes were then placed into a 1:1 solution of peroxide reagent and luminol enhancer reagent (Clarity max western ECL solution, Bio-Rad) for 30 seconds. Membranes were then imaged using a Bio-Rad ChemiDoc system using the automatic exposure feature to prevent over exposure of bands. Blots were stripped using stripping buffer (50mM glycine, 150NaCl, 0.1% Tween20, pH 2.5) for 20min before being re-probed with anti-actin antibodies overnight at 4°C. Antibodies used were anti-phospho-p42/44 MAPK (ERK) (Thr202/Tyr204) (1:1000, Cell Signaling), anti-phospho-Rac1/Cdc42 (Ser71) (1:1000, Cell Signaling), anti-beta actin (1:1,000, Abcam), anti-rabbit IgG HRP-conjugated (1:5,000, R&D Systems), anti-mouse IgG HRP-conjugated (1:5,000, R&D Systems).

### Reactive Oxygen Species Assays

Neutrophils (5×10^6^/ml) were incubated with either GM-CSF (5ng/ml), TNF-α (10ng/ml), or no treatment for 45min at 37°C with gentle rotation to induce cell priming. After 45min incubation, 2×10^5^ neutrophils were diluted into 200uL HBSS in a white 96 well plate (Corning Falcon® sterile microplate), stimulated with fMLP (1µM), PMA (100ng/ml), or a control of no stimulus along with luminol (10µM) and immediately placed into the BMG Labtech Fluorstar Optima plate reader. The luminometer was previously set to 37°C and luminol-enhanced chemiluminescence was recorded continuously for up to 30min.

### Chemotaxis Assays

Poly-Hema-coated 24-well plates were prepared and washed, before adding 800µL of media containing fMLP (10^-8^M, Sigma #F3506), IL-8 (100ng/mL, Sigma #I1645) or no stimulus. One hanging cell chamber with 3µm pores (Merck Millipore PTSP24H48) was placed into the media in each well and incubated at 37°C incubator for 10min before adding 200µL (10^6^) neutrophils. The plate placed in a 37°C incubator for 1.5h before chambers were removed. The media in each well was mixed via gentle pipetting before counting the number of migrated neutrophils.

### Flow Cytometry

Five hundred thousand neutrophils were stained with saturating concentrations of fluorescently-conjugated antibodies as previously described. Cells were fixed in 4% paraformaldehyde (PFA) before analysing on a Beckman Coulter CytoFLEX flow cytometer with a minimum of 10,000 events measured. Antibodies used were CD177 FITC Monoclonal Antibody (MEM-166, Thermo Fisher), CD54 (ICAM-1) PE-Vio615, REAfinity antibody (Miltenyi), CD181 (CXCR1) FITC REAfinity antibody (Miltenyi), CD182 (CXCR2) APC, REAfinity antibody (Miltenyi), REAfinity isotype controls (APC, PE-Vio615, FITC, Miltenyi) or unstained (US) controls. Fluorescence data were analysed using the software CytExpert v2.6.0.105.

### Neutrophil Extracellular Trap Assay

Neutrophils (5×10^6^) were incubated in phenol red-free RPMI 1640 media (with L-glutamine and HEPES) containing 5mL SYTOX Green Nucleic Acid Stain (Thermofisher) in a black 96 well plate (Greiner Bio one). Cells were allowed to settle for 15min prior to addition of LPS from *P. aeruginosa* (1 or 10ng/mL, Merck), PMA (0.1mg/mL, Merck), or A23187 (3.8μM, Merck) along with an untreated control. The plate was incubated for 4h at 37°C analysing on a Omega fluorescence plate reader with excitation at 480/490nm and read at 520nm.

### Neutrophil Extracellular Trap Microscopy

Neutrophils (2×10^5^) were incubated on poly-L-lysine-coated coverslips in a 24-well plate in 500uL RPMI media (+2% FCS) for 1h in CO_2_ 37°C. Following incubation, PMA (600nM), A23187 (3.8μM), LPS from *P. aeruginosa* (4ng/mL), or no stimulus were added and the plate incubated for 4h in CO_2_ at 37°C. Media was aspirated and cells were fixed with 200uL 4% PFA for 5min, before aspirating PFA and adding 500uL PBS for storing in the fridge at 4°C overnight. The next day, the glass cover slip was carefully removed using fine sterile forceps and stained as previously described [33]. Primary antibodies used were rabbit anti-neutrophil elastase and mouse anti-myeloperoxidase, each from Abcam used at 1:200 dilution). Secondary antibodies were anti-rabbit IgG alexafluor 488 and anti-mouse IgG alexafluor 647 (1:400-2000 dilution, Fisher scientific). DAPI stain solution was used at 1ug/and cells were washed and mounted onto glass slides using a drop (20uL) of mounting media (mowiol 488). Slides were imaged using a Zeiss LSM 800 confocal microscope at ×10 and X20. Images were processed using Zeiss Zen lite v3.11 software.

### Bacterial Killing Assay

Live *Staphylococcus aureus* (Oxford strain) were washed, suspended at 5L×L10^8^/ml in HBSS and opsonized with 10% human AB serum (Merck, Gillingham, UK) for 30min at 37°C. Neutrophils (5×10^6^/ml) were incubated with or without TNFα (10Lng/ml) and for up to 2h at 37°C with gentle agitation and serum-opsonized bacteria at a ratio of 1:10. Neutrophils were lysed to release live bacteria by serial dilution in distilled water and vigorous vortexing, before plating on LB agar and overnight incubation at 37°C. Bacteria colonies were counted the following day and the numbers from the treatment and no treatment control were compared with the colonies on the bacteria only control plate. Results were reported as a percentage of bacterial colonies killed by neutrophils compared to the bacteria only control.

### Serum Cytokine Arrays

Serum cytokine levels were quantified using the Proteome Profiler Human Cytokine Array Kit (R&D systems) following manufacturer’s protocol. Two sera were pooled per membrane (n=3 membranes per group) i.e. six sera in total were analysed across three membranes per disease group. Membranes were then placed and imaged in a Bio-Rad ChemiDoc imaging system. ImageJ (Fiji) was used to analyse the images using a macro to overlay the human cytokine array coordinates provided with the kit with the imaged spots. Membrane images were normalised to each other using the positive control reference spot intensities to establish an equal exposure scale for all values.

### Statistical Analysis of Functional Experiments

One-way ANOVA (performed in R v4.5.1) with Tukey’s HSD post-hoc tests was applied to determine the significant differences between groups. An adjusted p-value<0.05 was considered significant.

## Results

### Alterations in neutrophil gene expression during healthy ageing and ageing with frailty are characterised by inflammatory pathways and alterations in G-protein signalling

In order to measure the changes in neutrophil phenotype associated with healthy ageing and ageing with frailty, bulk RNAseq was performed on neutrophils from healthy younger people (HY, n=8), healthy older people (HO, n=9) and people with frailty (FR, n=10). A population of people with rheumatoid arthritis (RA, n=10) was also included to identify inflammatory signatures associated with ageing and/or frailty. Principal component analysis (PCA) of all samples together showed little separation between groups (**figure 1A**), with PC1 accounting for 21.7% of the variance of the transcriptional profiles of all four groups, while PC2 explained 10.2% of the variance. The HY and HO groups had a tighter clustering compared to the FR and RA groups. Among the most significant DEGs in FR neutrophils (**figure 1B**) was the matrix remodelling gene MXRA7 which was significantly elevated in FR neutrophils compared to HO (adj. p<0.01). Three genes associated with G-protein/GTPase signalling, RTP4, ADORA2A and RIN1, were significantly elevated in FR neutrophils compared to HO and HY neutrophils (adj. p<0.05). A summary of the number of up and down regulated genes between each group comparison is shown in **figure 1C**.

**Figure 1.**
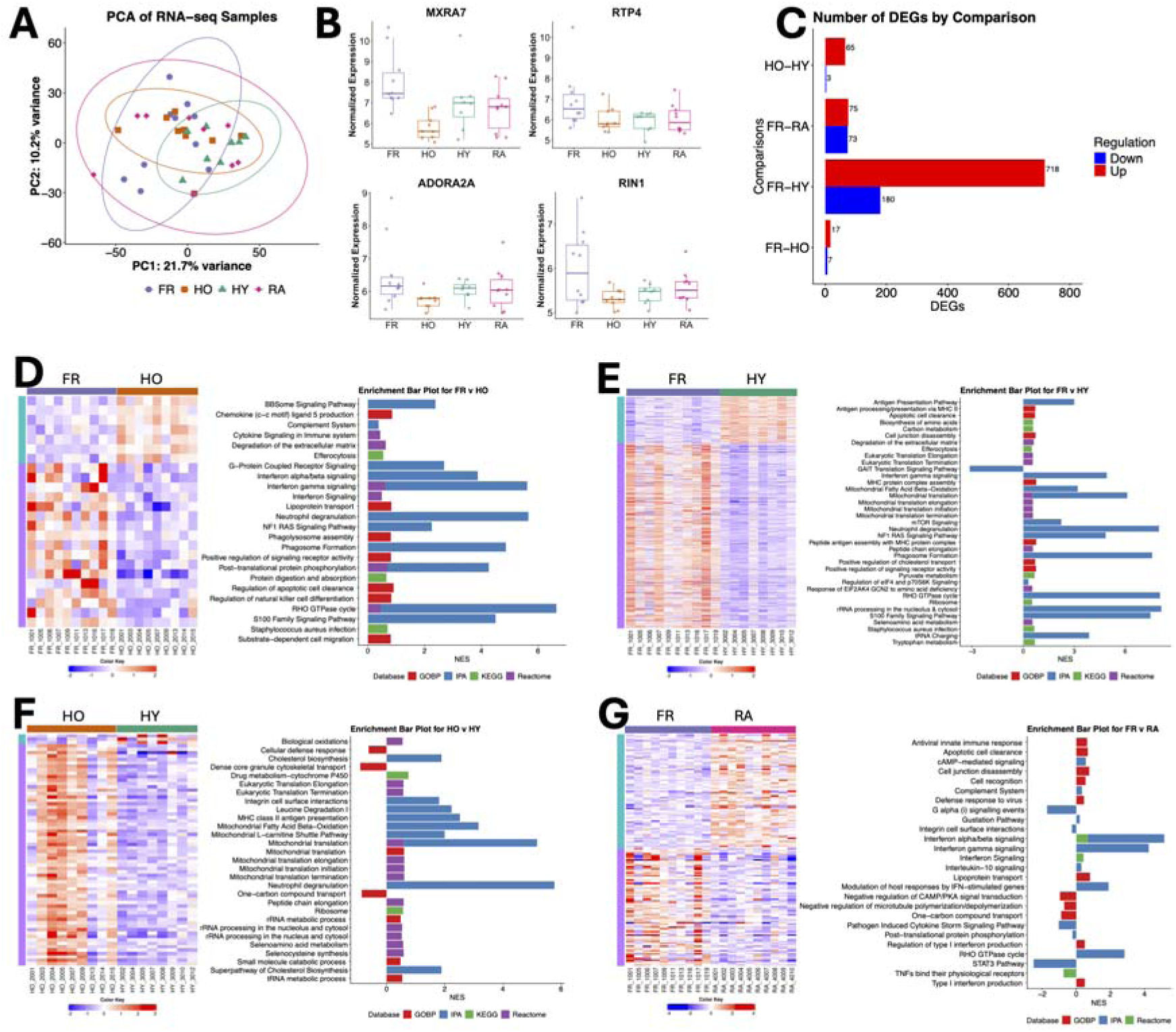
Transcriptomic analysis of neutrophils using RNAseq. (A) Principal component analysis (PCA) of neutrophil transcriptomes from frailty (FR, n=10, purple), healthy old (HO, n=9, orange), healthy young (HY, n=8, green) and rheumatoid arthritis (RA, n=10, pink). (B) Genes significantly elevated in FR neutrophils (adj. p<0.05). (C) Number of differentially expressed genes (DEGs) between comparisons (adj. p<0.05, fold change +/- 1.5). Heatmap of DEGs and pathway enrichment analysis for (D) FR vs HO, (E) FR v HY, (F) HO v HY, (G) FR v RA neutrophils. Heatmap (red = high, blue = low gene expression; purple bar = downregulated DEGs, blue bar = Upregulated DEGs). Expression analysis (NES = normalized enrichment score).

### Comparing Differences Between Healthy Old and Frailty

There were 24 significant DEGs between FR and HO (adj. *p*<0.05, < -1.5 or > 1.5-fold change). Of these, 17 were upregulated and 7 downregulated in FR neutrophils compared to HO as summarised by heatmap **(figure 1D).** Pathway enrichment of DEGs was performed via the web-based tool Integrated Differential Expression & Pathway analysis (IDEP) which incorporated various databases such as Gene Ontology Biological Processes (GOBP), KEGG, IPA and Reactome (**Supplementary table 2**). Immune-relevant pathways identified by enrichment analysis are shown in **figure 1D**. This included several pathways relating to G-Protein Coupled Receptor signalling and interferon signalling in FR neutrophils as well as pathways relevant to neutrophil activation such as cytokine signalling, neutrophil degranulation phagosome formation and S100 family protein signalling.

The GPRC signalling pathway was predicted to be upregulated in FR neutrophils, including predicted activation of ERK1/2 (**supplementary figure 1,** adj. p=5.23×10^-9^). Regulators of cytoskeleton remodelling such as RHOC, RHOU, ROCK1/2, PAK1 and PAK4 were expressed at higher levels in FR neutrophils (**supplementary figure 1**). FR neutrophils also had increased expression of GTPases that inhibit cytoskeleton remodelling not annotated in the IPA GPCR signalling pathway such as ARAP2 and ARHGAP24 indicating that cytoskeleton remodelling and directional sensing were dysregulated in neutrophils within frailty. Interferon signalling pathways were also enriched in FR compared to HO (**supplementary figure 2**). Overlaying of the FR vs HO gene expression data onto the IPA interferon signalling pathway showed significant upregulated of transcription factors (STAT1, STAT2) and genes in response to activation of both interferon alpha/beta receptors (IFIT1, OAS1, MX1, IFI6) and interferon gamma receptors (IFI35, **supplementary figure 2**). This indicated that there are inflammatory stimuli that have activated these pathways within FR neutrophils, which could indicate the chronic low-grade inflammation observed with inflammageing.

### Comparing Differences Between Frailty and Healthy Young

There were 898 significantly different expressed genes between FR and HY neutrophils (adj. *p*<0.05, (< -1.5 or > 1.5-fold change). Of these, 718 were upregulated and 180 were downregulated in FR compared to HY as summarised by heatmap (**figure 1E).** Pathway enrichment of DEGs in FR vs HY neutrophils was performed via IDEP and IPA (**Supplementary table 3**). Immune-relevant pathways enriched in FR neutrophils included antigen presentation via MHC class II pathways, eukaryotic translation pathways, mitochondrial translation pathways, neutrophil degranulation and phagosome formation which were elevated in FR neutrophils (**figure 1E**).

### Comparing Differences Between Healthy Old and Healthy Young

There were 68 significant DEGs between HO and HY neutrophils (adj. *p*<0.05, (< -1.5 or > 1.5-fold change). Of these, 65 were upregulated and 3 were downregulated in HO compared to HY as summarised by heatmap **(figure 1F).** Pathway enrichment of DEGs in HO vs HY neutrophils was performed via IDEP and IPA (**Supplementary table 4**). Immune-relevant pathways enriched in HO neutrophils included mitochondrial gene expression and metabolism, eukaryotic translation and ribosomal RNA (rRNA) processing, cholesterol biosynthesis, MHC class II antigen presentation and neutrophil degranulation. Cellular defence response, granule transport and one-carbon compound transport were predicted to be downregulated in HO neutrophils compared to HY **(figure 1F).**

### Comparing Differences Between Frailty and Rheumatoid Arthritis

There were 148 significantly different expressed genes between FR and RA neutrophils (adj. *p*<0.05, (< -1.5 or > 1.5-fold change). Of these, 75 were upregulated and 73 were downregulated in FR compared to HY as summarised by heatmap **figure 1G**. Pathway enrichment of DEGs in FR vs RA neutrophils was performed via IDEP and IPA (**Supplementary table 5**). Immune-relevant pathways enriched in FR neutrophils included interferon signalling pathways, Rho GTPase signalling, and complement signalling, whereas defence response, and STAT3 pathway were predicted to be downregulated in FR compared to RA, i.e. higher in RA neutrophils (**figure 1G**). Notably, IFN-stimulated gene expression was heterogeneous in RA neutrophils as previously reported [29, 34], and across the cohorts, IFN-stimulated genes were expressed at higher levels in FR neutrophils than in RA neutrophils (**supplementary figure 2).**

### Frailty is associated with elevated serum cytokines

Upstream regulator analysis was performed in IPA to predict which serum cytokines were activating neutrophils *in vivo* in health, RA and frailty based on the results of the RNAseq analysis. For comparison between FR and HO neutrophils, IPA predicted FR neutrophils were activated by a number of circulating cytokines, including IFNγ, TNFα, IL-1β and CSF2 (GM-CSF) (**figure 2A**). For HO and HY comparisons, cytokines IL-4, IL-5, CSF2 and TNFα were predicted to be activating HO neutrophils (**figure 2B**). For FR in comparison to HY, TNFα, IL-4, IFNγ, and CSF2 were activating FR neutrophils (**figure 2C**) whereas FR in comparison to RA predicted IFNL1 (IFNλ), IFNA2 (IFNα), IFNB1 (IFNβ) and IFNγ to be activating FR neutrophils more than RA neutrophils (**figure 2D**). Only IL1RN (IL1ra) was predicted to regulate RA neutrophils more than FR neutrophils. To validate these results, we measured the levels of 36 cytokines in serum from each participant group. FR sera had elevated levels of GM-CSF, IFN-γ, IL-1β, IL-8, CXCL1 and CXCL10 (**figure 2E**). Only CXCL10 was significantly higher in FR compared to HY via one way ANOVA and application of Tukey’s post-hoc tests (*p*<0.05). Removal of the RA group resulted in CXCL10 being significantly higher in FR than HO (*p*<0.05) and complement 5 (C5) being significantly lower in HY compared to HO (*p*<0.05) and FR (*p*<0.01, **figure 2F**). RA sera had elevated levels of IL-17α, IL-2, IL-4, IL-5, SerpinE1, and TNF-α (**figure 2G**) which correlates with previous literature on circulating cytokines within RA [35, 36]. These results confirm an inflammatory environment in frailty with some overlaps to the systemic inflammation seen in RA.

**Figure 2.**
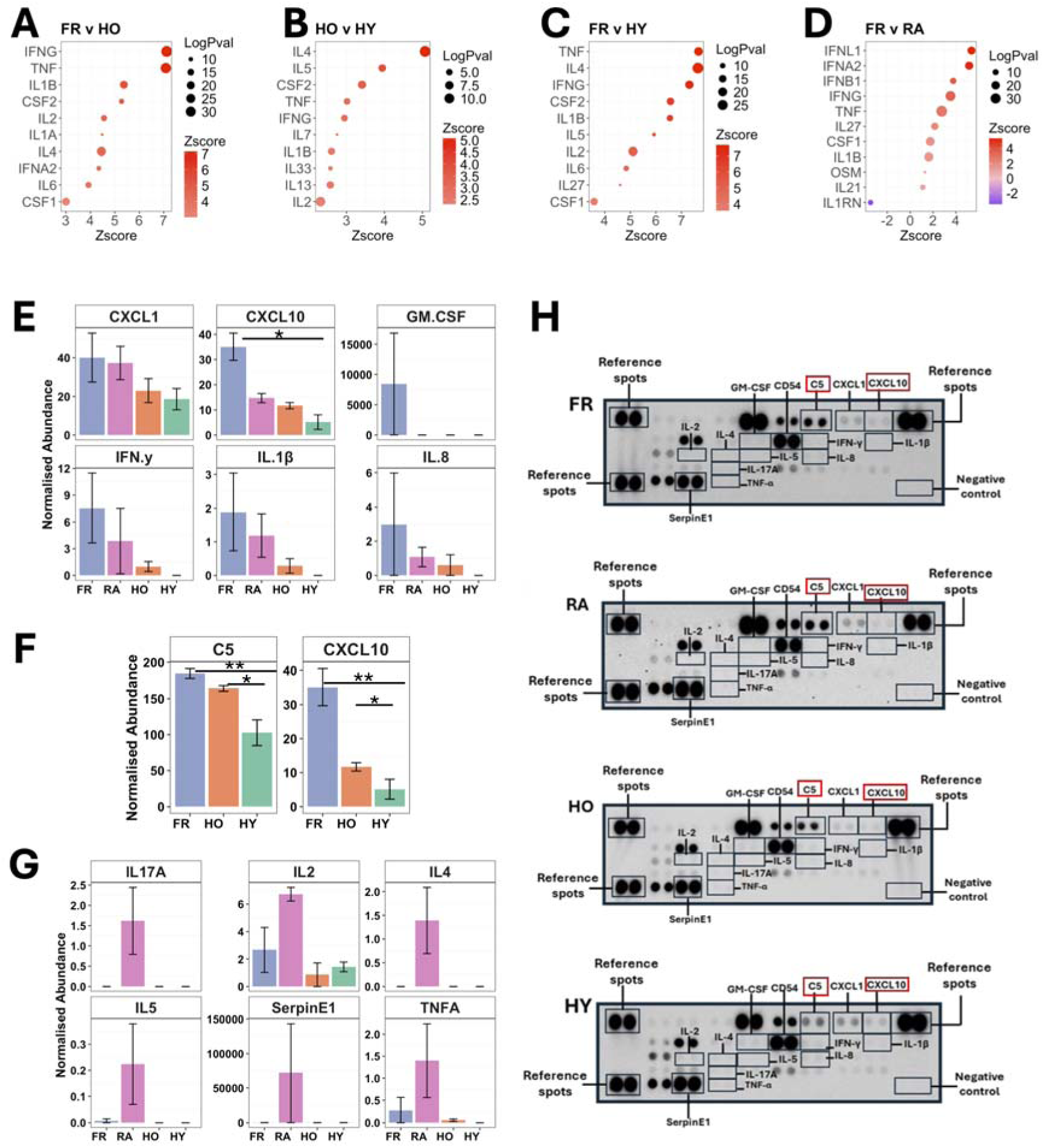
Cytokine activation of neutrophils in frailty. Predicted cytokine activators of neutrophil gene expression using IPA for (A) FR v HO, (B) HO v HY, (C) FR v HY, (D) FR v RA. (E) Cytokines in serum measured by protein array (FR, HO, HY, RA each n=6, *p<0.05 by ANOVA). (F) Excluding RA group, complement C5 and CXCL10 were significant by ANOVA (*p<0.05, ** p<0.01). (G) Cytokines elevated in RA sera. (H) Representative images of protein arrays with cytokine spots of interested highlighted (significant proteins in red boxes).

### Neutrophils from people with frailty have decreased chemotaxis ability

Functional experiments were subsequently performed to validate the transcriptomics analysis, which predicted inflammatory alterations to neutrophil phenotype associated with healthy ageing and ageing with frailty. We first wanted to investigate chemotactic ability of neutrophils in frailty, and whether we could detect the presence of reverse migrated neutrophils (ICAM1^high^/CXCR1^low^). We did by measuring the expression of both IL-8 chemokine receptors (CXCR1 and CXCR2), intercellular adhesion molecule-1 (ICAM-1, CD54), and CD177 on freshly isolated blood neutrophils using flow cytometry.

CXCR1 (CD181) and CXCR2 (CD182) expression showed no significant differences between HO, HY, FR and RA neutrophils via one-way ANOVA when RA was included in the analysis. The RA cohort appeared to skew the analysis due to its high donor-donor variability, heterogeneity in disease activity and wide age-range of participants (25-69 years) (**figure 3A**). Omitting the RA group from the analysis yielded significant differences between the healthy older and frail group for CXCR1 (adj. *p*<0.01) and CXCR2 (adj. *p*<0.00001), which were both higher in the frail group (**figure 3B**). Removal of RA also resulted in significant differences between HY and FR for CXCR1 (adj*. p*<0.0001), and CXCR2 (adj. *p*<0.00001) with both being higher in the FR group (**figure 3B**). There was no significant difference in CXCR1 and CXCR2 expression between HO and HY neutrophils. We found that CD177 expression was significantly elevated on FR neutrophils compared to HO (adj. *p*<0.05) and HY (adj. *p*<0.05), but there was no significant difference between HO and HY or FR and RA (**figure 3B**). This shows the differences are associated with inflammatory phenotype rather than age. We did not detect reverse migrated neutrophils (CD181+/CD182+/CD54+) in any participant group (data not shown).

**Figure 3.**
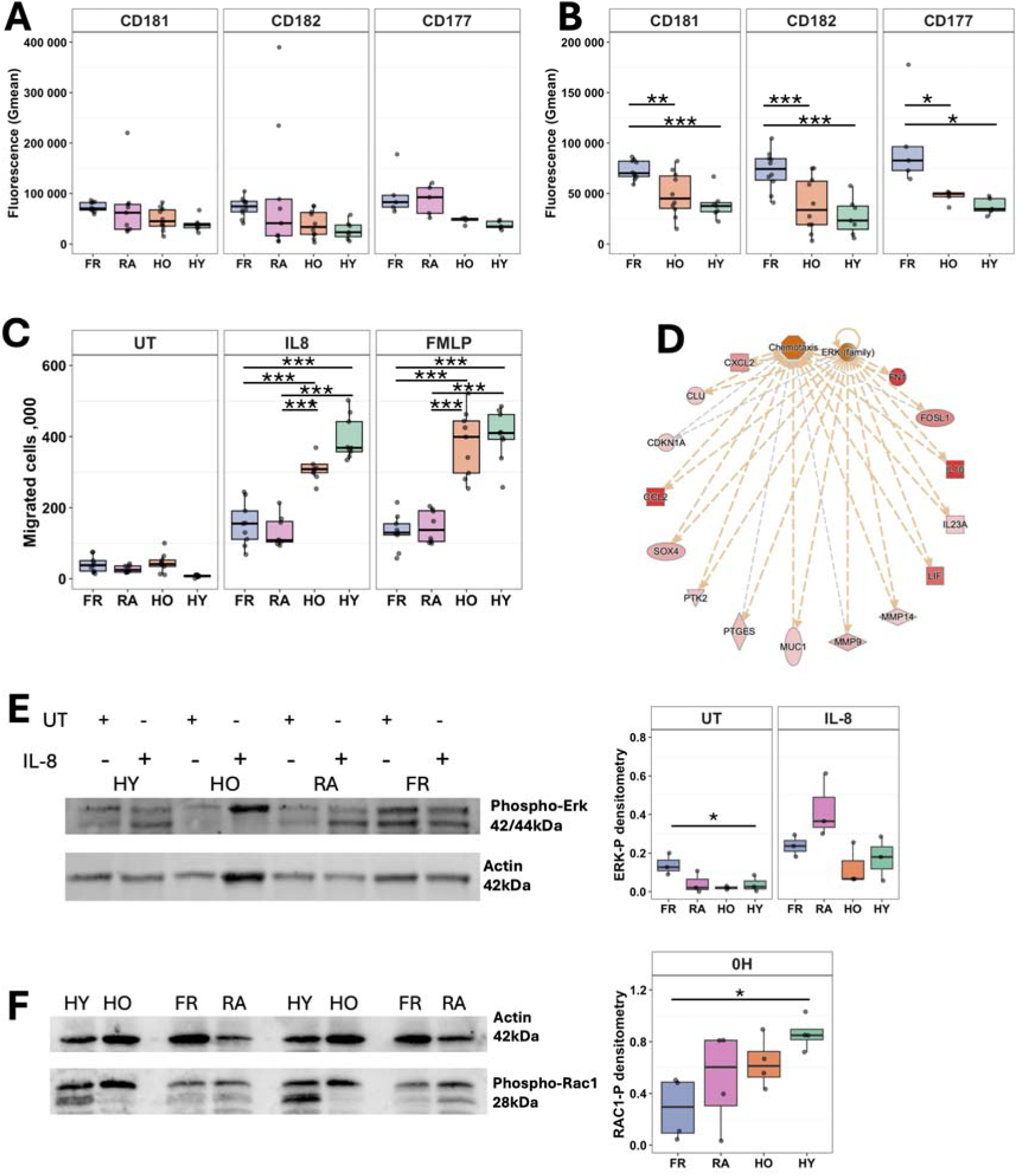
Impaired chemotaxis in frailty neutrophils. (A) Expression of CXCR1 (CD181), CXCR2 (CD182) and CD177 via flow cytometry. (B) Removal of the RA group resulted in significant differences. (C) Chemotaxis of neutrophils towards IL-8, fMLP or random migration (UT). (D) ERK-mediated signalling network predicted to be regulating chemotaxis in FR neutrophils (adj. p=5.23×10^-9^). Red = up-regulated gene expression, Orange = predicted activation. Representative western blots and densitometry (n=4) for (E) phosphorylated ERK (ERK-P) in untreated (UT) and 5 min IL-8 treated neutrophils and (F) phosphorylated RAC-1 (RAC1-P) in freshly isolated, 0h neutrophils. Purple = FR (n=4-10), Pink = RA (n=4-9), Orange = HO (n=4-10), Green = HY (n=4-7). Analysed by ANOVA (*p<0.05, **p<0.01, ***p<0.001).

Neutrophil migration towards IL-8, fMLP, or untreated media (random migration) was measured using a well-validated chemotaxis assay [37]. Significantly fewer FR neutrophils migrated towards IL-8 compared to HO neutrophils (adj. *p* < 0.001, **figure 3C**) and the same effect was observed with FR vs HO neutrophils migrating towards fMLP (adj. *p* < 0.001, **figure 3C**). There were also significantly fewer FR neutrophils migrating towards IL-8 (adj. *p* < 0.001, **figure 3C**), and fMLP (adj. *p* < 0.001, **figure 3C**) compared to HY neutrophils. HO neutrophils had lower rates of migration towards IL-8 and fMLP compared to HY neutrophils, but this was not statistically significant. There were no differences in migration in any treatment between FR and RA (p=0.99) and also no significant differences in random migration between all groups. These results indicate that whilst FR neutrophils express elevated IL-8 receptors on their plasma membrane, they have a reduced capacity to migrate towards IL-8 which may impair their ability to reach sites of infection *in vivo*.

### Altered ERK and Rac-1 activation is associated with dysfunctional chemotaxis in neutrophils from people with frailty

ERK1/2 was predicted by IPA as an upstream regulator of a network of genes regulating chemotaxis in FR neutrophils (**figure 3D**, adj. p=5.23×10^-9^). As IL-8 activates ERK signalling [38], neutrophils were either untreated or treated with IL-8 for 5 min and the levels of activated (phosphorylated) ERK (ERK-P) compared by Western blot. ERK-P was increased in HO, HY and RA neutrophils with IL-8 treatment. In FR neutrophils, ERK-P was significantly higher in the untreated cells but not in the treated cells compared to HO (*p*<0.05). ERK-P was also notably higher (although not statistically significant) in FR compared to HY untreated neutrophils (p=0.067), and no differences were observed between FR compared to RA, or HO compared to HY (**figure 3E**).

G-protein coupled receptor regulation of actin polymerisation was also predicted to be altered in FR neutrophils (**figure 1D**). PAK1 and PAK4 were predicted to be upregulated. Both these kinases are activated by RAC1, and therefore RAC1 phosphorylation was also measured in freshly isolated, 0h neutrophils. In FR neutrophils, RAC1-P was decreased compared to the other groups, with FR vs HY reaching statistical significance (p=0.03) (**figure 3F**).

### Neutrophils from people with frailty produce more ROS and NETs

Aberrant ROS production is a hallmark of neutrophil activation in inflammatory disease. In order to measure total ROS production, neutrophils were pre-incubated with GM-CSF, TNFα or no treatment for 45min, before being activated by fMLP (10^-3^M). ROS production was increased in FR neutrophils compared to HO (adj*. p*<0.05) and HY (adj. *p*<0.01) for the GM-CSF-primed neutrophils when stimulated with fMLP **(figure 4A)**. There were no differences between HO and HY or between FR and RA neutrophils. There were also no differences in ROS production when the neutrophils were unstimulated and untreated i.e. spontaneous ROS production (data not shown), and no differences between groups in ROS production when neutrophils were primed with TNFα **(figure 4A)**.

**Figure 4.**
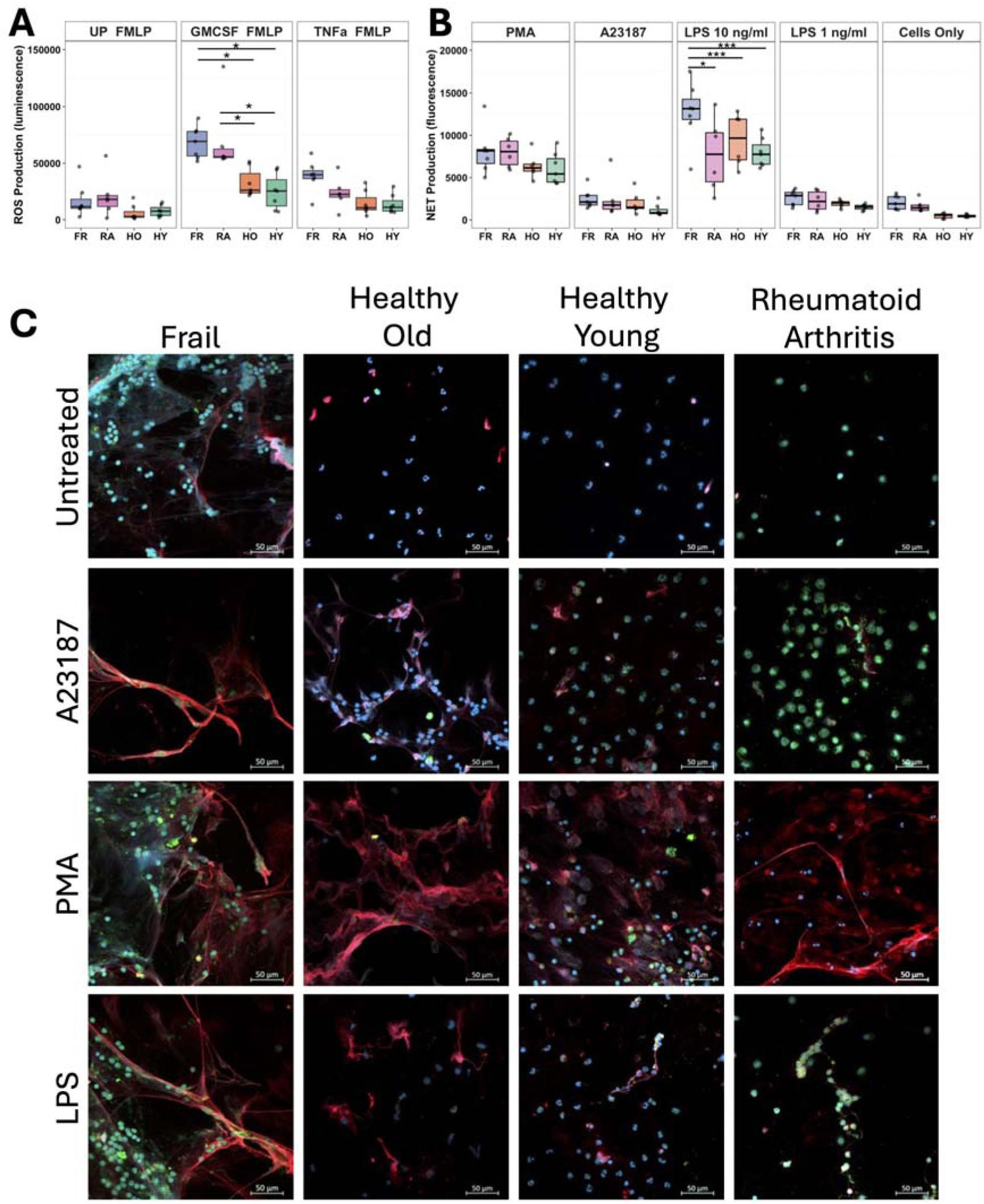
Neutrophil ROS and NET production in frailty. (A) ROS production in response to fMLP in neutrophils primed with GM-CSF (5ng/mL), TNFa (10ng/mL) or unprimed (UP). (B) NET production in response to PMA (0.1mg/mL), A23187 (3.8mM ), LPS (1 or 10 ng/mL) or unstimulated (cells only). Purple = FR (n=5-10), Pink = RA (n=5-9), Orange = HO (n=5-10), Green = HY (n=5-7). Analysis by ANOVA (*p<0.05, **p<0.01, ***p>0.001). (C) Representative microscopy images of NETs on coverslips (Blue = DNA, Red = MPO, Green = neutrophil elastase). Scale bar 50μM.

NET production was compared across all study participant groups in response to PMA (0.1mg/ml), A23187 (3.8mM) and LPS (10ng and 1ng /mL). The LPS used in these experiments was derived from *Pseudomonas aeruginos*a as it has been previously shown that neutrophils produce mixed responses to LPS from *E. coli* typically used in laboratory experiments [39]. NET production was significantly increased in FR neutrophils in response to LPS compared to HO (*adj. p*<0.0001), HY (*adj. p*<0.0001) and RA (*adj. p*<0.05, **figure 4B**). There was no significant difference in NET production in response to PMA and A23187 between the groups by ANOVA. The response to PMA was greater than A23187 across all groups, reflecting the difference between suicidal (PMA) and vital (A23187) NET production. Representative images of immunofluorescent-stained NETs are shown in **figure 4C**. Some spontaneous NET production was evident in untreated FR neutrophils. A23187-stimulated NETs covered a smaller area compared to PMA-stimulated NETs, which spread wider, covering a larger area. The response of HO, HY and RA neutrophils to LPS was similar to A23187, producing smaller NETs, whereas FR neutrophils produced much larger NETs that were visually similar to PMA-stimulated NETs. These results indicate that FR neutrophils have a greater capacity to produce ROS and NETs with the potential to cause localised inflammation and tissue damage.

### Neutrophils from people with frailty have reduced bacterial killing capacity

The bacterial killing capacity of neutrophils through ageing and with frailty was also investigated. Neutrophils were incubated with and without TNF-_α_ to induce priming, prior to addition of bacteria for 90min. Killing of live, opsonised *S. aureus* bacteria was significantly reduced in unprimed FR neutrophils compared to HO at 1hr (adj. *p*<0.05), HY at 1hr (adj. *p*<0.0001) and HY at 2 hr (adj. *p* <0.05, **figure 5**). Bacterial killing by untreated HO neutrophils was also significantly lower at 2h compared to HY (adj. *p* <0.05), and RA neutrophils had lower killing capacity at 1h and 2h compared to HY neutrophils (adj. *p* <0.05). For the TNF_α_ -primed neutrophils, there was a significant difference between FR and HY only after 2h (adj. *p* <0.05). There were no differences in killing capacity between FR and RA neutrophils. These results suggest that FR neutrophils have a reduced capacity to kill bacteria which correlates with reports of slower recovery from infection in people with frailty.

**Figure 5.**
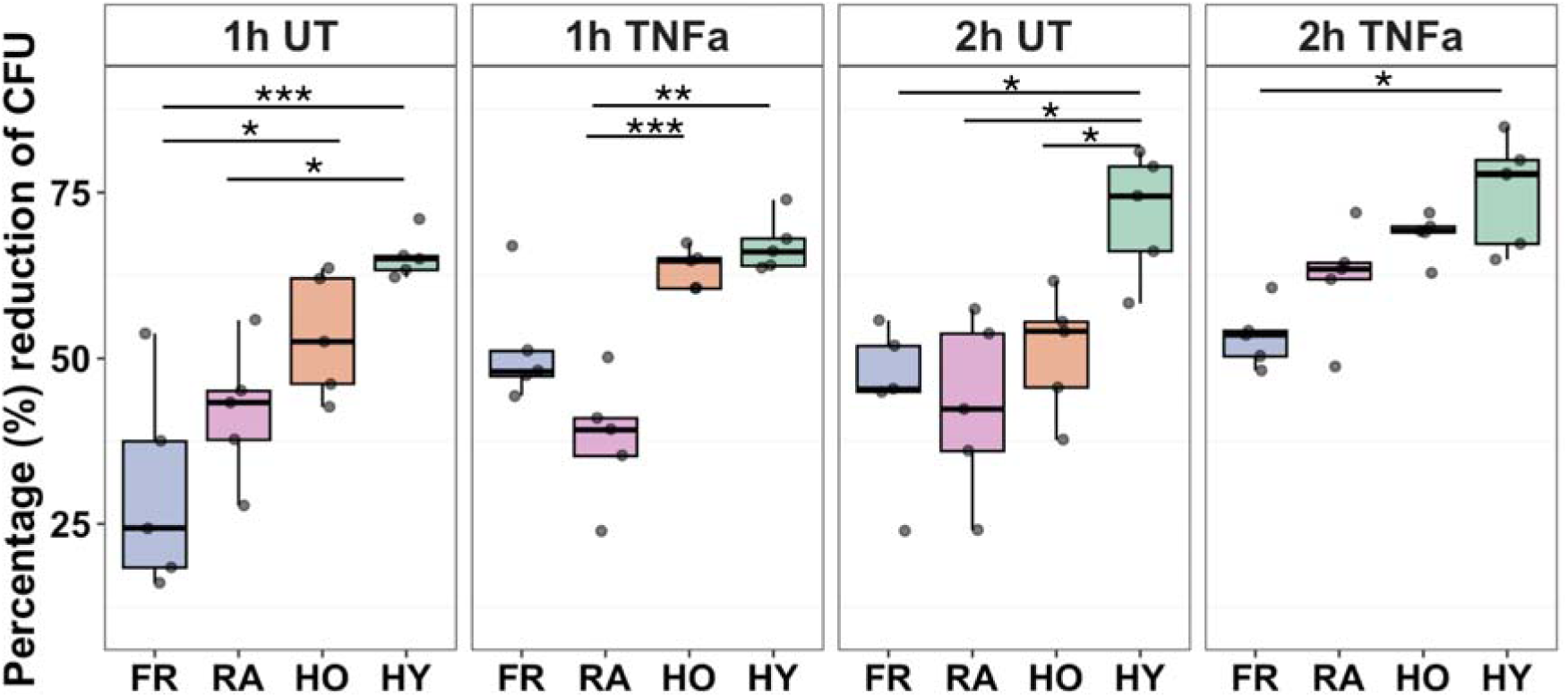
Neutrophil killing of *S. aureus* bacteria. Neutrophils were either untreated (UT) or primed with TNFα (10ng/mL) before incubating with *S. aureus* bacteria (10:1 ratio) for up to 2h. Purple = FR (n=5-10), Pink = RA (n=5-9), Orange = HO (n=5-10), Green = HY (n=5-7). Analysis by ANOVA (*p<0.05, **p<0.01, ***p>0.001).

## Discussion

In this study we have combined neutrophil transcriptomic analysis with functional experiments to describe an altered, activated neutrophil phenotype in frailty. Our bioinformatics analysis identified increased activation of immune-relevant pathways in FR neutrophils, including activation of interferon and cytokine receptor signalling, and activation of G-protein signalling regulated pathways as well as dysregulation in a number of metabolic pathways. We performed a number of functional experiments which showed that FR neutrophils had impaired chemotaxis to both IL-8 and fMLP compared to healthy neutrophils, along with increased production of ROS and NETs. Despite this activated phenotype, FR neutrophils had impaired bacterial killing capacity, suggesting an inflammageing phenotype. ERK signalling was also constitutively activated in FR neutrophils.

IL-8, also known as CXCL8, has two main receptors on neutrophils, CXCR1 (CD181) and CXCR2 (CD182), both of which are G-protein coupled receptors. It is known that the disruption of neutrophil recruitment to a site of infection could lead to mal containment of pathogens. This is supported by neutropenia being a significant risk factor for infection [40]. A primary function of IL-8 is to induce neutrophil recruitment towards a site of infection or injury. Neutrophil activation by IL-8 also upregulates adhesion molecules, increases intracellular calcium, and primes the neutrophil for oxidative burst [41, 42]. Our study showed that despite elevated expression of both IL-8 receptors on the cell surface, FR neutrophils had reduced migration towards IL-8 *in vitro*. Although there are no previous reports on IL-8 receptor expression within frailty, there are studies that looked at the changes in IL-8 receptor expression on neutrophils from older people compared to younger ages. These studies showed no differences in CXCR1 and CXCR2 expression between neutrophils from healthy young and healthy older subjects [43, 44]. There were also no differences in expression of CXCR1 or CXCR2 between older people who were active or unactive [45]. Although the frail group in this study had significantly higher expression of both IL-8 receptors, they had a significantly reduced chemotaxis towards IL-8 compared to the healthy groups. This could be due to a desensitisation of IL-8 within frail neutrophils, or a dysregulation of intracellular signalling mechanisms downstream of the IL-8 receptor, and ERK signalling was found to be constitutively activated in FR neutrophils. Serum analysis of cytokines identified elevated IL-8 levels in FR sera, suggesting FR neutrophils had already been exposed to IL-8 *in vivo*. Elevation of the chemokine CXCL1 from mast cells in an aged mouse model of inflamed tissue caused desensitisation of CXCR2 on neutrophils and loss of neutrophil directionality within endothelial cell junctions [46]. This caused them to reverse migrate out of the inflamed tissue and back into the circulation and cause vascular damage. This desensitisation could also be the case for neutrophils from people with frailty, wherein they express higher numbers IL-8 receptors to compensate for the lack of sensitivity.

As well as altered IL-8 receptor expression, a higher expression of CD177 was identified on FR neutrophils. While the exact function of CD177 is still poorly understood, it is known to be upregulated when stimulated via granulocyte colony stimulating factor (G-CSF), and during bacterial infections [47]. Antibody-mediated clustering of CD177 on the surface of neutrophils also primes for fMLP-activated respiratory burst [48]. Despite this, there are mixed reports on whether CD177 expression is increased on neutrophils in sites of inflammation. CD177+ neutrophils are found increased in inflamed tissues such as within periodontitis [49], and in inflammatory bowel disease (IBD) [50]. There are also reports showing no differences between CD177+ and CD177- neutrophils in tissues such as the peritoneum [51]. However, systemically there is evidence of increased CD177 in septic shock and in ICU patients with Covid-19 [52, 53] and levels of CD177 discriminated between recovery and death of patients with Covid-19 [53]. CD177 expression is associated with certain autoimmune diseases such as ANCA-associated vasculitis [54] and RA [55]. Within RA specifically, patients taking methotrexate (MTX) had lower CD177 expression compared to MTX-naive patients [55]. In patients with IBD, *in vitro* experiments showed CD177+ neutrophils released more ROS, MPO and NETs, but lower levels of proinflammatory cytokines (e.g., IL-6 and IFN-γ) than CD177- neutrophils [56]. There is a debate as to whether CD177+ aids in neutrophil transmigration. One study showed that that CD177 promotes neutrophil migration via PECAM-1 [57]. A more recent study showed ligation of CD177+ neutrophils on endothelial cells reduced transmigration *in vitro* via a PECAM-1-independent pathway [58]. There are no specific studies looking at whether CD177 increases with age and/or frailty status, however, the wider literature seems to indicate that it is increased by inflammatory conditions rather than by age.

Chemotaxis and neutrophil migration deficits have been described in aged hosts compared to young hosts. Neutrophils isolated from older subjects exhibit markedly reduced migration in response to the bacterial products fMLP and LPS [59]. However, there has been only one study which looks at chemotaxis within neutrophils from people with frailty [6]. Similarly to our findings, this study showed that chemotactic deficits were exacerbated within frailty compared to healthy older adults [6]. The addition of young plasma to old neutrophils could not reverse the age-associated migratory phenotype [44], and exposure of young neutrophils to plasma from aged hosts did not mimic the ageing-related decline in chemotaxis [44]. Our Western blot analysis showed that ERK is constitutively activated in FR. Despite this, FR neutrophils had reduced RAC-1 expression. RAC-1 controls directionality within neutrophils, with RAC-1 deficient mice developing multiple leading protrusions of the actin cytoskeleton instead of one, directed leading edge towards the chemoattractant gradient [60]. RAC-1 activates PI3K, leading to PIP_3_ production and accumulation at the leading edge of the migrating cell [61]. Depletion or overexpression of PI3K negatively affects chemotaxis [62]. Two studies have shown that inhibition of PI3K signalling improved chemotaxis in healthy older adults and older adults with frailty towards IL-8 [6, 44]. A PI3K knockout model was still able to migrate with near wild-type efficiency but had clear defects in migration speed [63], whereas chemical inhibition of PI3K caused delays in neutrophil migration speed towards fMLP [64, 65].

FR neutrophils had significant upregulation in the pathways involved in inflammation compared to HO. These pathways included interferon signalling, cytokine signalling, neutrophil degranulation, phagosome formation and *S. aureus* infection. Type I IFN signalling (IFNα/IFNβ) was previously identified as being elevated in a sub-set of people with RA [29, 34], so an increase in IFN signalling in FR neutrophils above that seen in RA neutrophils is intriguing. Activation of the IFN signalling pathway can also lead to cellular senescence which increases with age [50] [66]. Additionally, autoimmunity to type I IFNs is a strong and common predictor of Covid-19 death [67, 68]. This increase in autoimmunity could be one of the many ways in which the frail immune system declines. Previous studies have found that IFN-γ inhibits random chemotaxis and regulates directional migration in human neutrophils [69]. Thus increased IFN-γ signalling could be driving the impaired chemotaxis phenotype seen in FR neutrophils in this study. The identification of upregulated degranulation pathways in FR compared to HO and HY (but not RA) supports the theory that neutrophils are activated within frailty in the absence of an appropriate stimulus. Degranulation is the result of downstream signalling events that begin with Toll-Like Receptor, GPCR, Fc Receptor or integrin receptor signalling pathways. The toxic effects of degranulation are not pathogen specific and damage healthy cells and tissues. This has been observed in autoimmune diseases including RA in which the neutrophils degranulate into the cartilage within the joints [70]. Degranulation products such as MPO, NE, MMPs, and cytokines degrade the cartilage and increase the pro-inflammatory response toward the joint.

This study clearly demonstrated that neutrophils from people with frailty are capable of producing ROS, and when primed with GM-CSF and stimulated by fMLP, they produce significantly more ROS compared to the two healthy groups, with no differences compared to RA. RA neutrophils have been shown to be primed for ROS production *in vivo* within sterile environments [71]. This is also supported by the increased levels of cytokines in RA sera such as TNFα and IL-1β, shown in the cytokine array blots, which can activate neutrophils. FR sera had significantly higher CXCL10 compared to both HO and HY. CXCL10, also known as interferon gamma-induced protein 10 (IP-10), is a chemokine that is produced in response to IFN-γ. Neutralisation of CXCL10 ameliorated the severity of ARDS by limiting neutrophil influx into the lung [72]. Activated neutrophils also produce CXCL10 in order to recruit T-cells to a site of inflammation [73]. It is unclear whether increased neutrophil counts cause an increase in circulating CXCL10 or whether increased CXCL10 causes neutrophil activation in frailty. Complement C5 was also significantly higher in FR and HO sera compared to the HY group. It is known that complement protein fragments (e.g. C5a) can activate neutrophils [74]. These circulating serum cytokines represent a way in which blood neutrophils could be activated *in vivo* and, therefore, primed for ROS production *in vitro*. Since they are already primed, FR and RA neutrophils are able to produce large amounts of ROS when stimulated *in vitro*, compared to neutrophils from healthy controls [75]. RA neutrophils are primed due to the circulating immune mediators such as G-CSF, IL-8, TNFα, and LTB_4_ [76]. It is known that people with frailty have an increased level of circulating pro-inflammatory molecules such as CRP, interleukin-1β (IL-1β) and interleukin-6 (IL-6) and TNFα compared to non-frail age matched controls [77], confirmed by the cytokine array in this study. This is due to the increase of senescent cells releasing pro-inflammatory cytokines.

A previous study on neutrophil ROS production showed that neutrophils from people with frailty produced significantly higher ROS in response to *Streptococcus pneumoniae* bacteria compared to younger adults but did not include a healthy older group for an age group comparison [78]. Indeed, there are currently a lack of studies comparing the ROS production of frail neutrophils to non-frail age matched controls. Higher levels of inflammation including PMA-induced oxidative burst were also reported in whole blood assays of older people with a slow gait speed (<0.8m/s) compared to those with a high gait speed (>0.8m/s) [79]. Although not explicitly clinically frail, slow gait speed is a physical manifestation of frailty. There were no significant differences in spontaneous ROS production in this study, however previous studies have shown that spontaneous ROS is higher in neutrophils from older compared to younger control groups [80] [81] [43]. It is worth noting that these studies did not separate their cohorts into healthy older and frail older groups, with exclusion criteria for active or inflammatory diseases yet no reports on the frailty status of the older participants.

This study also identified an increase in NETs production by frail neutrophils when stimulated with LPS from *P. aeruginosa.* Previous studies have shown greater production of NETs in elderly subjects (over 65 years) than in younger adults (under 50 years), but these NETs also differed in structure and size, with the NETs from the elderly group being larger [82]. This is in contrast to a previous study in neutrophils from aged mice showed significantly fewer NETs in response to *S. aureus in vitro* and *in vivo* [3]. Our data show that neutrophils from humans with frailty are still able to produce NETs and their ability is not affected by age or frailty status. Currently there are no other studies investigating NET production in neutrophils from people with frailty and without an active infection. Despite this, there are studies from different infections in the elderly population that correlate with our data. Covid-19 hospitalisations in people aged over 65 showed that a 3x increase of neutrophil elastase within serum of deceased patients compared to those who survived, and was an independent predictor of death [83]. Within sarcopenia, spontaneous NET formation was significantly elevated, with no differences in NET formation in response to *E. coli* [84]. It is thought that neutrophils developed the ability to produce NETs in response to pathogens based on their size. For example, bacteria can be engulfed and phagocytosed, however, large filamentous fungi would require exogenous methods of elimination [85]. Despite this, it is known that neutrophils do also produce NETs in response to bacteria. Although NETs can aid in degradation of bacteria, they can cause harm as in the case of bacterial keratitis from *Pseudomonas aeruginosa* infection. In this case in mice, neutrophils produce NETs to contain the bacterial biofilms to the external corneal environment and prevent dissemination into the brain but causes blindness as a result [86]. In the bacterial keratitis model, when NETs were blocked via knockout, the *P. aeruginosa* biofilm broke down, reversed the keratitis, but allowed the bacteria to enter brain via the corneal surface, leading to extreme brain infection [86].

One key result from this study is that initially (within the first hour) the bacterial killing capacity of FR neutrophils was impaired compared to the HO group. However bacterial killing capacity after 2h was not significantly different. This could indicate that FR neutrophils are slower to phagocytose and/or release cytotoxic molecules into the phagosome to kill bacteria. Evidence of bacterial killing capacity in people of different ages and within frailty is scarce, with only limited studies investigating bacterial killing capacity using neutrophils from inflammatory diseases and animal studies. Many pro-inflammatory diseases present with dysregulated neutrophil functions. One example is a recent study where people from the ages of 30-75 were separated into groups based on whether they had obesity, diabetes, metabolic syndrome, or healthy controls, however they were not stratified by age. There were no differences between the groups in chemotaxis towards multiple stimuli including fMLP and IL-8. Additionally, there were no differences for phagocytosis-mediated killing of *S. aureus* [87]. It is possible that the mixed age ranges within groups was causing the lack of significant differences, and that the groups would benefit from further separation by their age. In a mouse model of obesity, insulin treatment improved neutrophil phagocytic and bactericidal activity towards *S. aureus* within the surgical site infection [88]. In a model of septic peritonitis (inflammation of the abdominal wall), older mice had a reduced capacity to clear pathogenic *E. coli* compared to young mice [89]. This included an impairment of NET release in response to *S. aureus* that was independent of LPS stimulation. Unexpectedly, chemotaxis was normal within the old mice [89]. Neutrophils from old mice failed to effectively kill *S. pneumoniae* even when the bacteria were opsonized with immune sera from young controls [90]. Bacterial killing of *S. aureus* and *P. aeruginosa* was significantly reduced in mild and severe Covid-19 patients compared to healthy controls regardless of age (participants were between 21-83 years old) [91]. This shows that regardless of age, neutrophils have a reduced bacterial killing capacity during infections which is a key factor driving secondary infection development.

We acknowledge some limitations in our study design. Firstly, bulk RNA sequencing was performed on a mixed population of freshly isolated blood neutrophils. There is limited evidence that frailty has an increased number of immature neutrophils which are not as functionally sophisticated as mature neutrophils [78]. Previous studies have shown that the circulating pool of healthy blood neutrophils contains sub-populations of neutrophils at different stages of maturity, with differential activation of transcription factors and different gene expression profiles. These changes are also evident during inflammation [92]. Single-cell RNA sequencing on neutrophils from people with frailty could identify differing stages of maturity and effector functions. Increasing the participant size for each group would also allow for more distinct differences to emerge between and within groups, specifically for the healthy older and healthy younger group.

Neutrophils from people with RA were included as an inflammatory control group in our study. However, it should also be noted that RA group had a wide age range as it included anyone above 18 years old with diagnosed RA. The oldest person with RA in this study was 69 and the youngest was 25 years old. The large range of ages in the RA group could have introduced variation in the neutrophil transcriptomes induced by ageing rather than disease activity that is unaccounted for. Therefore, to know whether these changes are solely due to disease or age, future studies should include an age-matched RA group to compare with the FR group. This would allow for more precise differences to be observed between FR and RA. All blood samples for this study were taken between 8am and 11am, with neutrophil isolations being performed within 1h of blood draw. However, the effects of circadian transcriptomic differences could still affect the results. Previous studies have shown that “freshly released” (from the bone marrow) neutrophils produce more NETs as opposed to “aged” neutrophils that had been in circulation for longer [93]. In this case “aged” neutrophils refers to the neutrophils that have spent a longer time in circulation and are homing back to the bone marrow for clearance, not to be confused with neutrophils from “aged people”. Although neutrophils follow a circadian rhythm where their numbers peak in the afternoon/evening, it would be interesting to know if any differences in their transcriptome occurred between the early hours of the morning (8am) compared to the later hours of the morning (11am).

Finally, in terms of functional experiments in this study, our conclusions are limited by our sample size which range from 5-10 participants per group. With a larger sample size, more differences may be distinguished between the groups and provide more robust evidence for the impaired neutrophil functions found within frailty. Only one timepoint was measured for ROS, NETs and chemotaxis. With more timepoints included in future work, it could reveal valuable information, for example, the frail neutrophils might be slower than the HO neutrophils but still reach the stimulus in the same numbers, albeit after longer periods of time. The bacterial killing assay was performed only with *S. aureus* bacteria as this is a well optimised assay of human neutrophil bacterial killing capacity. In future studies, the bacterial killing assay could be extended to include relevant pathogens such as *P. aeruginosa* and *E.coli* to further examine the killing capacity of FR neutrophils.

In summary, this study has demonstrated that the neutrophil transcriptome diverges with age, and in healthy ageing compared to ageing with frailty. Our analyses identified impaired migration capacity in frailty neutrophils linked to a dysregulation of G-protein regulated genes. Additionally, although FR and RA neutrophils have both derived from low-grade inflammatory environments, they displayed significant transcriptomic differences showing different upstream regulators of activation in both phenotypes. An emerging model is that FR neutrophils are activated *in vivo,* likely due to the increased levels of chronic inflammation (inflammageing) compared to HO neutrophils. We showed that although activated, FR neutrophils have a reduced capacity to migrate towards inflammatory stimuli and a reduced capacity to kill live bacteria. This could be due to circulating inflammatory signals acting a as false stimulus, and/or defects in regulation of signalling pathway activation. Due to this altered phenotype, FR neutrophils may be causing harm to healthy tissues, either through impaired migration, or increased NETs and ROS production. Identifying the specific pathways which are altered in frailty could lead to therapeutics that restore a normal neutrophil phenotype. This area of neutrophil research is key for future drug development and drug repurposing, as exaggerated and/or aberrant neutrophil activity can be detrimental to tissues. Frailty research is relatively new and a focus should be placed on determining the differences that cause neutrophils to become dysregulated. This is important as people with immune frailty contract more infections and have a higher mortality rate from infections compared to non-frail older adults [94–96]. If it is determined that similar inflammatory pathways are regulated in frailty and in RA, this opens the possibility for drug repurposing from rheumatology into the management of people with frailty.

## Ethics Statement

The study was approved by the NRES Southwest Central Bristol Research Ethics Committee for the collection of blood serum from healthy controls and people with frailty (Ref: 21/SW/0093). All participants gave written, informed consent in accordance with the declaration of Helsinki.

## Data Availability

All data produced in the present study are available upon reasonable request to the authors

## Acknowledgements

We would like to thank the clinical staff at Liverpool University Hospital NHS Foundation Trust for help recruiting people with frailty and RA for this study. We would also like to thank Urszula Poplawska for technical support with microscopy imaging.

## Authorship Declaration

G.A.A. performed the experiments, formal analysis, methodology, visualisation, writing – original draft. J.W. performed the experiments, visualisation, writing – review and editing. A.S. performed the experiments, writing – review and editing. C.M. performed the experiments, formal analysis, writing – review and editing. A.A. supervision, participant screening and recruitment, writing – review and editing. M.M.P. conceptualisation, supervision, funding acquisition, visualisation, writing – original draft. H.L.W. conceptualisation, funding acquisition, methodology, data curation, formal analysis, project administration, supervision, visualisation, writing – original draft.

