## Supplementary Data for "Altered neutrophil G-protein receptor signalling linked to impaired chemotaxis and increased ROS and NET production in older people with frailty"

**^2^Institute of Systems, Molecular and Integrative Biology, University of Liverpool, Liverpool UK**

**3School of Biosciences, University of Liverpool UK**

^4^Bunbury Regional Hospital, Bunbury, Western Australia

^5^Division of Internal Medicine, University of Western Australia, Perth, Western Australia

**^6^High-Field NMR Facility, Liverpool Shared Research Facilities (LivSRF) University of Liverpool, Liverpool UK**

**Supplementary Table 1.** Demographics of study participants for neutrophil transcriptomics analysis. FR = people with frailty, HO = healthy older people, HY = healthy younger people, RA = people with rhwumatoid arthritis.

|  | **FR** | **HO** | **HY** | **RA** |
| --- | --- | --- | --- | --- |
| **Average age** | 82 | 70 | 22 | 53 |
| **Age matching pval** | FR:HO  *p*<0.05 | FR:HY  *p*<0.05 | HO:HY  *p*<0.05 | FR:RA  *p*<0.05 |
| **% Female** | 60 | 40 | 75 | 90 |
| **n** | 10 | 9 | 8 | 10 |

**Supplementary Table 2.** IDEP and IPA enrichment analysis of FR v HO neutrophil gene expression. NES = normalised enrichment score, GOBP = gene ontology biological process, KEGG = Kyoto Encyclopedia of Genes and Genomes, IPA = Ingenuity Pathway Analysis, Reactome = Reactome Pathway Database.

| **Pathway** | **NES** | **P value** | **- Log10 P value** | **Database** |
| --- | --- | --- | --- | --- |
| Negative regulation of retinoic acid receptor signaling pathway | -0.9215 | 1.0e-02 | 2.00 | GOBP |
| Positive regulation of signaling receptor activity | 0.8157 | 2.3e-03 | 2.64 | GOBP |
| Negative regulation of oxidative stress-induced neuron intrinsic apoptotic signaling pathway | 0.9041 | 6.9e-03 | 2.16 | GOBP |
| Lipoprotein transport | 0.8276 | 1.0e-02 | 2.00 | GOBP |
| Lipoprotein localization | 0.8135 | 1.4e-02 | 1.85 | GOBP |
| Regulation of basement membrane organization | 0.9011 | 1.5e-02 | 1.82 | GOBP |
| Phagolysosome assembly | 0.8119 | 1.5e-02 | 1.82 | GOBP |
| Delamination | 0.9528 | 1.7e-02 | 1.77 | GOBP |
| Regulation of apoptotic cell clearance | 0.9068 | 1.8e-02 | 1.74 | GOBP |
| Regulation of neurotransmitter receptor activity | 0.8073 | 1.8e-02 | 1.74 | GOBP |
| Substrate-dependent cell migration | 0.8059 | 1.8e-02 | 1.74 | GOBP |
| Amyloid-beta clearance by transcytosis | 0.9042 | 2.0e-02 | 1.70 | GOBP |
| Neuron intrinsic apoptotic signaling pathway in response to oxidative stress | 0.7998 | 2.4e-02 | 1.62 | GOBP |
| Lymphangiogenesis | 0.8829 | 2.9e-02 | 1.54 | GOBP |
| Chemokine (c-c motif) ligand 5 production | 0.8543 | 3.1e-02 | 1.51 | GOBP |
| Regulation of chemokine (c-c motif) ligand 5 production | 0.8543 | 3.1e-02 | 1.51 | GOBP |
| Regulation of nmda receptor activity | 0.8636 | 3.4e-02 | 1.47 | GOBP |
| Synapse pruning | 0.8451 | 3.6e-02 | 1.44 | GOBP |
| Regulation of natural killer cell differentiation | 0.8257 | 4.3e-02 | 1.37 | GOBP |
| Regulation of oxidative stress-induced neuron intrinsic apoptotic signaling pathway | 0.8005 | 4.7e-02 | 1.33 | GOBP |
| Systemic lupus erythematosus | 0.7691 | 1.5e-02 | 1.82 | KEGG |
| Amoebiasis | 0.615 | 3.4e-02 | 1.47 | KEGG |
| Efferocytosis | 0.5445 | 3.4e-02 | 1.47 | KEGG |
| Staphylococcus aureus infection | 0.6955 | 4.8e-02 | 1.32 | KEGG |
| Protein digestion and absorption | 0.655 | 4.8e-02 | 1.32 | KEGG |
| Telomere C-strand Lagging Strand Synthesis | -0.619 | 1.5e-02 | 1.82 | Reactome |
| Regulation of Insulin-like Growth Factor IGF transport and uptake by Insulin-like Growth Factor Binding Proteins IGFBPs | 0.6963 | 5.8e-04 | 3.24 | Reactome |
| Post-translational protein phosphorylation | 0.6843 | 4.7e-03 | 2.33 | Reactome |
| Extracellular matrix organization | 0.5522 | 6.3e-03 | 2.20 | Reactome |
| Immune System | 0.3889 | 8.1e-03 | 2.09 | Reactome |
| Cytokine Signaling in Immune system | 0.4352 | 8.4e-03 | 2.08 | Reactome |
| Binding and Uptake of Ligands by Scavenger Receptors | 0.7656 | 1.2e-02 | 1.92 | Reactome |
| Degradation of the extracellular matrix | 0.629 | 1.2e-02 | 1.92 | Reactome |
| Interferon gamma signaling | 0.6172 | 1.2e-02 | 1.92 | Reactome |
| RHO GTPase cycle | 0.4536 | 1.5e-02 | 1.82 | Reactome |
| Interferon Signaling | 0.4896 | 2.5e-02 | 1.60 | Reactome |
| Interferon gamma signaling | 5 | 2.51e-11 | 10.60 | IPA |
| Phagosome Formation | 4.866 | 1.45e-08 | 7.84 | IPA |
| S100 Family Signaling Pathway | 4.503 | 9.55e-08 | 7.02 | IPA |
| BBSome Signaling Pathway | 2.38 | 2.95e-07 | 6.53 | IPA |
| NF1 RAS Signaling Pathway | 2.252 | 4.79e-07 | 6.32 | IPA |
| Post-translational protein phosphorylation | 3.578 | 6.76e-07 | 6.17 | IPA |
| Neutrophil degranulation | 5.657 | 3.02e-06 | 5.52 | IPA |
| Interferon alpha/beta signaling | 3.873 | 9.55e-06 | 5.02 | IPA |
| RHO GTPase cycle | 6.193 | 1.35e-05 | 4.87 | IPA |
| G-Protein Coupled Receptor Signaling | 2.689 | 4.07e-05 | 4.39 | IPA |
| Complement System | 0.378 | 1.51e-04 | 3.82 | IPA |

**Supplementary Table 3.** IDEP and IPA enrichment analysis of FR v HY neutrophil gene expression. NES = normalised enrichment score, GOBP = gene ontology biological process, KEGG = Kyoto Encyclopedia of Genes and Genomes, IPA = Ingenuity Pathway Analysis, Reactome = Reactome Pathway Database.

| **Pathway** | **NES** | **P value** | **- Log10 P value** | **Database** |
| --- | --- | --- | --- | --- |
| Muscle organ morphogenesis | 0.6963 | 2.1e-03 | 2.68 | GOBP |
| Apoptotic cell clearance | 0.693 | 5.8e-03 | 2.24 | GOBP |
| Antigen processing and presentation of peptide or polysaccharide antigen via mhc class ii | 0.7103 | 7.6e-03 | 2.12 | GOBP |
| Collagen catabolic process | 0.7681 | 1.3e-02 | 1.89 | GOBP |
| Positive regulation of signaling receptor activity | 0.7532 | 1.4e-02 | 1.85 | GOBP |
| Positive regulation of synaptic transmission glutamatergic | 0.7855 | 1.8e-02 | 1.74 | GOBP |
| Muscle tissue morphogenesis | 0.672 | 1.8e-02 | 1.74 | GOBP |
| Synapse pruning | 0.8507 | 2.4e-02 | 1.62 | GOBP |
| Regulation of neurotransmitter receptor activity | 0.7732 | 3.4e-02 | 1.47 | GOBP |
| Regulation of vascular associated smooth muscle cell differentiation | 0.8088 | 3.7e-02 | 1.43 | GOBP |
| Negative regulation of transforming growth factor beta production | 0.9134 | 4.1e-02 | 1.39 | GOBP |
| Negative regulation of oxidative stress-induced neuron intrinsic apoptotic signaling pathway | 0.8573 | 4.1e-02 | 1.39 | GOBP |
| Renal filtration cell differentiation | 0.7963 | 4.3e-02 | 1.37 | GOBP |
| Podocyte differentiation | 0.7963 | 4.3e-02 | 1.37 | GOBP |
| Glomerular epithelial cell differentiation | 0.7963 | 4.3e-02 | 1.37 | GOBP |
| Mhc protein complex assembly | 0.7621 | 4.3e-02 | 1.37 | GOBP |
| Peptide antigen assembly with mhc protein complex | 0.7621 | 4.3e-02 | 1.37 | GOBP |
| Cell junction disassembly | 0.7222 | 4.3e-02 | 1.37 | GOBP |
| Positive regulation of sterol transport | 0.7029 | 4.3e-02 | 1.37 | GOBP |
| Positive regulation of cholesterol transport | 0.7029 | 4.3e-02 | 1.37 | GOBP |
| Systemic lupus erythematosus | 0.7734 | 2.2e-03 | 2.66 | KEGG |
| Carbon metabolism | 0.5799 | 3.9e-03 | 2.41 | KEGG |
| Metabolic pathways | 0.3987 | 3.9e-03 | 2.41 | KEGG |
| Ribosome | 0.5324 | 5.4e-03 | 2.27 | KEGG |
| Pyruvate metabolism | 0.6674 | 3.0e-02 | 1.52 | KEGG |
| Staphylococcus aureus infection | 0.6518 | 3.0e-02 | 1.52 | KEGG |
| Efferocytosis | 0.5057 | 3.0e-02 | 1.52 | KEGG |
| Tryptophan metabolism | 0.6862 | 3.2e-02 | 1.49 | KEGG |
| Biosynthesis of amino acids | 0.5764 | 3.2e-02 | 1.49 | KEGG |
| Intestinal immune network for IgA production | 0.6581 | 4.2e-02 | 1.38 | KEGG |
| Biosynthesis of cofactors | 0.5059 | 4.2e-02 | 1.38 | KEGG |
| Binding and Uptake of Ligands by Scavenger Receptors | 0.7577 | 5.7e-03 | 2.24 | Reactome |
| Degradation of the extracellular matrix | 0.6201 | 5.7e-03 | 2.24 | Reactome |
| Mitochondrial translation termination | 0.5713 | 1.2e-02 | 1.92 | Reactome |
| Peptide chain elongation | 0.5709 | 1.2e-02 | 1.92 | Reactome |
| Mitochondrial translation elongation | 0.5674 | 1.2e-02 | 1.92 | Reactome |
| Eukaryotic Translation Elongation | 0.566 | 1.2e-02 | 1.92 | Reactome |
| Eukaryotic Translation Termination | 0.5598 | 1.2e-02 | 1.92 | Reactome |
| Nonsense Mediated Decay NMD independent of the Exon Junction Complex EJC | 0.5556 | 1.2e-02 | 1.92 | Reactome |
| Selenocysteine synthesis | 0.5532 | 1.2e-02 | 1.92 | Reactome |
| Formation of a pool of free 40S subunits | 0.551 | 1.2e-02 | 1.92 | Reactome |
| Mitochondrial translation | 0.5484 | 1.2e-02 | 1.92 | Reactome |
| Mitochondrial translation initiation | 0.5646 | 1.4e-02 | 1.85 | Reactome |
| Viral mRNA Translation | 0.5641 | 1.4e-02 | 1.85 | Reactome |
| Scavenging of heme from plasma | 0.8582 | 2.5e-02 | 1.60 | Reactome |
| Selenoamino acid metabolism | 0.5444 | 2.7e-02 | 1.57 | Reactome |
| Response of EIF2AK4 GCN2 to amino acid deficiency | 0.5339 | 2.7e-02 | 1.57 | Reactome |
| L13a-mediated translational silencing of Ceruloplasmin expression | 0.5248 | 2.7e-02 | 1.57 | Reactome |
| PD-1 signaling | 0.764 | 3.4e-02 | 1.47 | Reactome |
| Drug ADME | 0.6081 | 4.5e-02 | 1.35 | Reactome |
| Regulation of Insulin-like Growth Factor IGF transport and uptake by Insulin-like Growth Factor Binding Proteins IGFBPs | 0.5747 | 4.5e-02 | 1.35 | Reactome |
| rRNA processing in the nucleolus & cytosol | 8.124 | 1.58e-11 | 10.80 | IPA |
| RHO GTPase cycle | 8.045 | 3.55e-10 | 9.45 | IPA |
| Neutrophil degranulation | 7.985 | 1.82e-08 | 7.74 | IPA |
| Mitochondrial translation | 5.568 | 4.37e-07 | 6.36 | IPA |
| tRNA Charging | 3.873 | 7.94e-07 | 6.10 | IPA |
| Interferon gamma signaling | 4.914 | 1.15e-06 | 5.94 | IPA |
| Antigen Presentation Pathway | 3 | 5.50e-06 | 5.26 | IPA |
| Mitochondrial Fatty Acid Beta-Oxidation | 3.207 | 1.70e-05 | 4.77 | IPA |
| Regulation of eIF4 and p70S6K Signaling | 0.302 | 1.78e-05 | 4.75 | IPA |
| S100 Family Signaling Pathway | 7.488 | 1.82e-05 | 4.74 | IPA |
| GAIT Translation Signaling Pathway | -3.157 | 2.09e-05 | 4.68 | IPA |
| mTOR Signaling | 2.236 | 2.24e-05 | 4.65 | IPA |
| NF1 RAS Signaling Pathway | 4.849 | 2.63e-05 | 4.58 | IPA |
| Phagosome Formation | 7.587 | 3.16e-05 | 4.50 | IPA |

**Supplementary Table 4.** IDEP and IPA enrichment analysis of HO v HY neutrophil gene expression. NES = normalised enrichment score, GOBP = gene ontology biological process, KEGG = Kyoto Encyclopedia of Genes and Genomes, IPA = Ingenuity Pathway Analysis, Reactome = Reactome Pathway Database.

| **Pathway** | **NES** | **P value** | **- Log10 P value** | **Database** |
| --- | --- | --- | --- | --- |
| Cellular defense response | -0.6149 | 8.80e-03 | 2.06 | GOBP |
| One-carbon compound transport | -0.8558 | 1.70e-02 | 1.77 | GOBP |
| Axonal transport | -0.5529 | 2.40e-02 | 1.62 | GOBP |
| Dense core granule cytoskeletal transport | -0.9057 | 2.90e-02 | 1.54 | GOBP |
| Dense core granule transport | -0.9057 | 2.90e-02 | 1.54 | GOBP |
| Axo-dendritic transport | -0.4936 | 2.90e-02 | 1.54 | GOBP |
| Carbon dioxide transport | -0.9273 | 4.40e-02 | 1.36 | GOBP |
| Mitochondrial translation | 0.6013 | 4.90e-04 | 3.31 | GOBP |
| Mitochondrial gene expression | 0.5735 | 4.90e-04 | 3.31 | GOBP |
| Ventricular cardiac muscle cell action potential | 0.8506 | 2.90e-03 | 2.54 | GOBP |
| Trna metabolic process | 0.5402 | 2.90e-03 | 2.54 | GOBP |
| Small molecule biosynthetic process | 0.4688 | 2.90e-03 | 2.54 | GOBP |
| Action potential | 0.6176 | 9.00e-03 | 2.05 | GOBP |
| Cardiac muscle cell action potential involved in contraction | 0.7492 | 2.30e-02 | 1.64 | GOBP |
| Small molecule catabolic process | 0.4787 | 2.30e-02 | 1.64 | GOBP |
| Carboxylic acid biosynthetic process | 0.4956 | 2.40e-02 | 1.62 | GOBP |
| Cardiac muscle cell action potential | 0.6718 | 2.90e-02 | 1.54 | GOBP |
| Organic acid biosynthetic process | 0.4939 | 2.90e-02 | 1.54 | GOBP |
| Rrna metabolic process | 0.4865 | 2.90e-02 | 1.54 | GOBP |
| Rrna processing | 0.4971 | 3.40e-02 | 1.47 | GOBP |
| R-HSA-76009 Platelet Aggregation Plug Formation | -0.6018 | 3.70e-02 | 1.43 | Reactome |
| R-HSA-5368286 Mitochondrial translation initiation | 0.6126 | 4.90Ee-03 | 2.31 | Reactome |
| R-HSA-5389840 Mitochondrial translation elongation | 0.6073 | 4.90e-03 | 2.31 | Reactome |
| R-HSA-5419276 Mitochondrial translation termination | 0.6059 | 4.90e-03 | 2.31 | Reactome |
| R-HSA-6791226 Major pathway of rRNA processing in the nucleolus and cytosol | 0.5363 | 4.90e-03 | 2.31 | Reactome |
| R-HSA-8868773 rRNA processing in the nucleus and cytosol | 0.524 | 4.90e-03 | 2.31 | Reactome |
| R-HSA-5368287 Mitochondrial translation | 0.5928 | 6.50e-03 | 2.19 | Reactome |
| R-HSA-2408557 Selenocysteine synthesis | 0.5893 | 7.90e-03 | 2.10 | Reactome |
| R-HSA-156902 Peptide chain elongation | 0.5887 | 8.40e-03 | 2.08 | Reactome |
| R-HSA-192823 Viral mRNA Translation | 0.5887 | 8.40e-03 | 2.08 | Reactome |
| R-HSA-156842 Eukaryotic Translation Elongation | 0.5845 | 8.40e-03 | 2.08 | Reactome |
| R-HSA-2408522 Selenoamino acid metabolism | 0.5718 | 8.40e-03 | 2.08 | Reactome |
| R-HSA-72764 Eukaryotic Translation Termination | 0.5764 | 1.10e-02 | 1.96 | Reactome |
| R-HSA-211859 Biological oxidations | 0.5515 | 1.80e-02 | 1.74 | Reactome |
| R-HSA-72689 Formation of a pool of free 40S subunits | 0.5551 | 2.20e-02 | 1.66 | Reactome |
| R-HSA-156588 Glucuronidation | 0.9047 | 3.50e-02 | 1.46 | Reactome |
| R-HSA-391908 Prostanoid ligand receptors | 0.8908 | 3.50e-02 | 1.46 | Reactome |
| R-HSA-975956 Nonsense Mediated Decay NMD independent of the Exon Junction Complex EJC | 0.5559 | 3.60e-02 | 1.44 | Reactome |
| R-HSA-168273 Influenza Viral RNA Transcription and Replication | 0.5122 | 3.70e-02 | 1.43 | Reactome |
| R-HSA-391903 Eicosanoid ligand-binding receptors | 0.8092 | 4.90e-02 | 1.31 | Reactome |
| Metabolic pathways | 0.4278 | 4.30e-05 | 4.37 | KEGG |
| Ribosome | 0.5679 | 3.10e-03 | 2.51 | KEGG |
| Drug metabolism-cytochrome P450 | 0.7533 | 3.30e-02 | 1.48 | KEGG |
| Mitochondrial translation | 4.583 | 5.37e-06 | 5.27 | IPA |
| Mitochondrial Fatty Acid Beta-Oxidation | 3.162 | 7.59e-05 | 4.12 | IPA |
| Leucine Degradation I | 2.236 | 1.07e-04 | 3.97 | IPA |
| Integrin cell surface interactions | 1.807 | 2.88e-04 | 3.54 | IPA |
| MHC class II antigen presentation | 2.524 | 4.90e-04 | 3.31 | IPA |
| Superpathway of Cholesterol Biosynthesis | 1.89 | 5.62e-04 | 3.25 | IPA |
| Mitochondrial L-carnitine Shuttle Pathway | 2 | 8.71e-04 | 3.06 | IPA |
| Cholesterol biosynthesis | 1.89 | 1.20e-03 | 2.92 | IPA |
| Neutrophil degranulation | 5.774 | 1.29e-03 | 2.89 | IPA |

**Supplementary Table 5.** IDEP and IPA enrichment analysis of FR v RA neutrophil gene expression. NES = normalised enrichment score, GOBP = gene ontology biological process, KEGG = Kyoto Encyclopedia of Genes and Genomes, IPA = Ingenuity Pathway Analysis, Reactome = Reactome Pathway Database.

| **Pathway** | **NES** | **P value** | **- Log10 P value** | **Database** |
| --- | --- | --- | --- | --- |
| Oxygen transport | -0.9247 | 9.60e-03 | 2.02 | GOBP |
| One-carbon compound transport | -0.8879 | 1.50e-02 | 1.82 | GOBP |
| Negative regulation of retinoic acid receptor signaling pathway | -0.9572 | 1.70e-02 | 1.77 | GOBP |
| Gas transport | -0.8893 | 2.60e-02 | 1.59 | GOBP |
| Negative regulation of microtubule polymerization or depolymerization | -0.7123 | 3.80e-02 | 1.42 | GOBP |
| Carbon dioxide transport | -0.942 | 4.10e-02 | 1.39 | GOBP |
| B cell receptor signaling pathway | -0.5947 | 4.10e-02 | 1.39 | GOBP |
| Regulation of camp/pka signal transduction | -0.9566 | 4.80e-02 | 1.32 | GOBP |
| Negative regulation of camp/pka signal transduction | -0.9566 | 4.80e-02 | 1.32 | GOBP |
| Positive regulation of stem cell proliferation | -0.7445 | 4.80e-02 | 1.32 | GOBP |
| Defense response to virus | 0.4504 | 4.90e-03 | 2.31 | GOBP |
| Apoptotic cell clearance | 0.6755 | 2.10e-02 | 1.68 | GOBP |
| Antiviral innate immune response | 0.6364 | 3.70e-02 | 1.43 | GOBP |
| Coronary vasculature development | 0.7175 | 4.00e-02 | 1.40 | GOBP |
| Synapse pruning | 0.868 | 4.10e-02 | 1.39 | GOBP |
| Cell junction disassembly | 0.7575 | 4.10e-02 | 1.39 | GOBP |
| Regulation of type i interferon production | 0.4975 | 4.10e-02 | 1.39 | GOBP |
| Type i interferon production | 0.4975 | 4.10e-02 | 1.39 | GOBP |
| Lipoprotein transport | 0.8041 | 4.80e-02 | 1.32 | GOBP |
| Cell recognition | 0.5428 | 4.80e-02 | 1.32 | GOBP |
| Hemostasis | -0.3749 | 7.50e-03 | 2.12 | Reactome |
| Developmental Biology | -0.3253 | 1.50e-02 | 1.82 | Reactome |
| TNFs bind their physiological receptors | -0.7498 | 3.20e-02 | 1.49 | Reactome |
| Interferon alpha/beta signaling | 0.6825 | 2.60e-04 | 3.59 | Reactome |
| Interferon Signaling | 0.4242 | 3.40e-02 | 1.47 | Reactome |
| Interferon alpha/beta signaling | 4.472 | 1.35e-08 | 7.87 | IPA |
| cAMP-mediated signaling | 0.557 | 7.94e-07 | 6.1 | IPA |
| Gustation Pathway | 0.2 | 7.24e-06 | 5.14 | IPA |
| Complement System | 0.333 | 1.05e-05 | 4.98 | IPA |
| G alpha (i) signalling events | -1.715 | 1.07e-05 | 4.97 | IPA |
| RHO GTPase cycle | 2.828 | 1.35e-05 | 4.87 | IPA |
| Post-translational protein phosphorylation | -0.229 | 1.48e-05 | 4.83 | IPA |
| Interferon gamma signaling | 4.243 | 1.55e-05 | 4.81 | IPA |
| Pathogen Induced Cytokine Storm Signaling Pathway | -1.029 | 3.89e-05 | 4.41 | IPA |
| Interleukin-10 signaling | 0.302 | 4.57e-05 | 4.34 | IPA |
| Modulation of host responses by IFN-stimulated genes | 1.89 | 6.61e-05 | 4.18 | IPA |
| STAT3 Pathway | -2.5 | 6.76e-05 | 4.17 | IPA |
| Integrin cell surface interactions | -0.258 | 1.23e-04 | 3.91 | IPA |

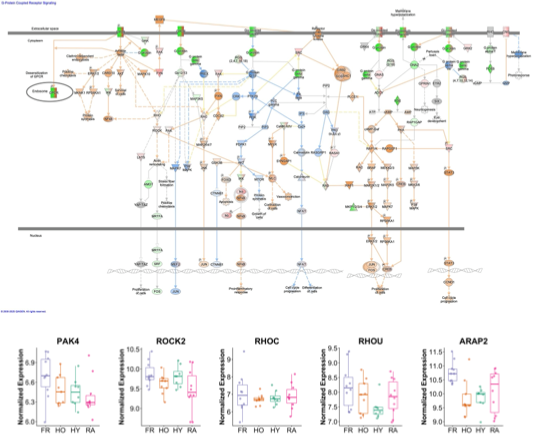

**Supplementary Figure 1. IPA G-protein coupled receptor pathway overlaid with FR v HO gene expression data.** Red = upregulated, Green = downregulated, Orange = predicted increase, Blue = predicted decrease. Box plots show normalized expression of key pathway genes significantly elevated in FR neutrophils.

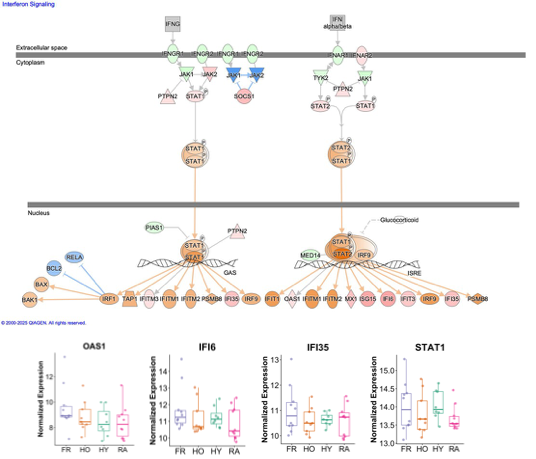

**Supplementary Figure 2. IPA interferon pathway overlaid with FR v HO gene expression data.** Red = upregulated, Green = downregulated, Orange = predicted increase, Blue = predicted decrease. Box plots show normalized expression of key pathway genes significantly elevated in FR neutrophils.
